# The Optimal Allocation of Covid-19 Vaccines

**DOI:** 10.1101/2020.07.22.20160143

**Authors:** Ana Babus, Sanmay Das, SangMok Lee

**Affiliations:** Department of Economics, Washington University in St. Louis; Department of Computer Science, George Mason University

**Keywords:** optimal assignment, occupational health, H4, I1

## Abstract

Covid-19 vaccine prioritization is key if the initial supply of the vaccine is limited. A consensus is emerging to first prioritize populations facing a high risk of severe illness in high-exposure occupations. The challenge is assigning priorities next among high-risk populations in low-exposure occupations and those that are young and healthy but work in high-exposure occupations. We estimate occupation-based infection risks and use age-based infection fatality rates in a model to assign priorities over populations with different occupations and ages. Among others, we find that 50-year-old food-processing workers and 60-year-old financial advisors are equally prioritized. Our model suggests a vaccine distribution that emphasizes age-based mortality risk more than occupation-based exposure risk. Designating some occupations as essential does not affect the optimal vaccine allocation unless a stay-at-home order is also in effect. Even with vaccines allocated optimally, 7.14% of the employed workforce is still expected to be infected with the virus until the vaccine becomes widely available, provided the vaccine is 50% effective, and assuming a supply of 60mil doses.

## 1 Introduction

The ongoing severe acute respiratory syndrome–coronavirus 2 (SARS-CoV-2) pandemic has claimed over 1,100,000 lives worldwide as of October 20th, 2020, and paralyzed economic activity around the globe for extended periods. Vaccination is seen as the principal strategy for containing the pandemic. However, even though vaccines are being developed at historical speed, they are expected to be initially in limited supply (Cohen, 2020). A crucial question stands: who will get first access to a vaccine when one becomes available?

While there is little disagreement that high-risk individuals in high-exposure occupations, such as front-line healthcare workers, should be included in the initial priority group for vaccination (National Academies of Sciences, 2020), assigning priorities over other groups remains subject to debate. To start with, a disproportionate toll on the elderly suggests that age should be the primary consideration. At the same time, infection clusters arising in hospitals and meatpacking plants indicate that there are occupations in which the risk of exposure to the virus is substantially raised. Deciding whether to prioritize meatpacking plant workers over elderly citizens must factor in the ages of the workers and the occupations of the elderly.

Moreover, the coronavirus pandemic is unique in its wide-spread impact on economic activity. Non-pharmaceutical interventions, such as social distancing and stay-at-home orders, have been implemented on an unprecedented scale. While many countries have relaxed constraints, lockdowns are expected to be imposed again as the virus re-surges. Thus, vaccines perhaps should be allocated not only based on the risk of infection or death but also based on the economic benefits of allowing certain groups of people to work earlier than others.

We develop and estimate a model to evaluate vaccination allocation strategies. We recognize that people face different levels of infection risks depending on their occupations and that, conditional on being infected, the risk of death depends on their ages. The vaccine is assumed to be effective only to some extent and in limited supply relative to the entire population. A vaccine distribution strategy may be supplemented by a targeted stay-at-home order that prevents certain age-occupation groups from returning to their workplaces at an economical cost.

We solve a simple linear program that takes into account the cost that an individual expects to incur if infected and the economic benefit from going back to her workplace. Our procedure allows us to derive the optimal vaccine distribution among all allocations based on occupation and age. This enables us to address who should get the vaccine earlier than others: young meatpacking plant workers or elderly school teachers?

To assign priorities over populations with different occupations and ages, we estimate occupation-based exposure risks (i.e., infection rates), and use estimates of age-based infection mortality rates. The infection mortality rates vary across ages far more than the estimated infection rates across occupations. Accordingly, our model suggests a vaccine distribution that emphasizes age-based mortality risk more than occupation-based exposure risk. This insight is robust to supplementing the vaccine distribution with a stay-at-home mandate for targeted occupation and age groups. If we consider a specification in which some occupations can be done from home, then the vaccine can be distributed to younger individuals who need to return to their workplaces. However, if the supply of the vaccine is scarcer, occupation-based exposure risks become more relevant as we distribute vaccines to individuals in relatively lower-risk occupations only at very advanced ages.

Our model implies that, even when vaccines are allocated optimally, 7.14% of the employed workforce is still expected to be infected until the vaccine becomes widely available, provided the vaccine is 50% effective and assuming a supply of 60mil doses. Either increasing the effectiveness of the vaccine or increasing the vaccine supply while keeping the effectiveness constant decreases the proportion of people infected with the virus. To curb the coronavirus-related deaths to a level comparable to seasonal flu the vaccine must be at least 65% effective and a much more stringent stay-at-home order is inevitable, even when vaccines are allocated optimally.

The allocation of vaccines has also been analyzed in the context of SEIR modeling framework (Bubar et al., 2020, Chen et al., 2020, Matrajit et al., 2020). However, these papers propose allocations solely based on age. Pathak et al. (2020) also discusses the allocation of scarce resources, including vaccines, during a pandemic. While we focus on which groups to prioritize, Pathak et al. (2020) studies the implementation of a given proportional prioritization (i.e. vaccine reserves for different groups). To illustrate, if the population is partitioned in healthcare vs. non-healthcare workers, and old vs. young, our model assigns priorities among young healthcare workers and elderly non-healthcare workers, based on occupation-related exposure and age-based infection mortality. In contrast, Pathak et al. (2020) takes a certain priority structure, in the form of reserves for, e.g., healthcare workers and the elderly, as given. Their primary question is how to implement a distribution of vaccines under the given priorities.^1^

## 2 The Model

We develop a simple model to identify priority groups for vaccination based on occupations and ages. For this, we partition the population into groups by occupations *I* and ages *J*. The population distribution over occupations and age-groups is 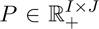 such that each element *p*_*ij*_ denotes the number of people with certain occupation *i* and in an age-group *j*. Each person in occupation *i* faces a risk of infection denoted by *r*_*i*_. Associated with each person in an age-group *j* is a cost of infection *c*_*j*_. Implicit here is the assumption that all population is susceptible to infection and could benefit from receiving a vaccine. We believe that the population that is immune to the virus is probably small and little is known about the duration of immunity.

A policy consists of a distribution of a limited supply of vaccines and a targeted stay-at-home order. A vaccine distribution 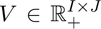 specifies the number of people, *v*_*ij*_, in occupation *i* and age-group *j* that receive a vaccine. The vaccine distribution satisfies a supply-side budget constraint ∑_*i,j*_ *v*_*ij*_ *≤ b*, where *b* represents the quantity of vaccine initially available, and it is assumed to be less than the total population ∑_*i,j*_*p*_*ij*_. The vaccine allocation policy can be supplemented by a targeted stay-at-home order 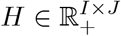 with *h*_*ij*_ representing the number of people in occupation *i* and age-group *j* that cannot return to their workplace. Overall, *V + H ≤ P*.

A vaccine recipient becomes immune to the virus with probability *γ*, which captures the vaccine’s effectiveness. It is worth noting that the vaccine in our model either prevents infections or reduces the risk of severe illness (captured by the cost of infection). Thus, each dose of the vaccine allocated to group *(i, j)* reduces *r*_*i*_*c*_*j*_ by the effectiveness rate, *γ*. Then, a vaccination policy, *V*, together with a stay-at-home order, *H*, can decrease the population affected by the virus by γ*V + H* across different occupations and age groups. In particular, for each group *(i, j)*, the policy saves costs (γ*v*_*ij*_ *+ h*_*ij*_*)r*_*i*_*c*_*j*_.

The stay-at-home policy *H* comes with the suspension of economic activities. Let 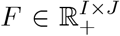 denote the values of economic activities that accrue to individuals. In particular, each *f*_*ij*_ denotes the value of the economic activity *i* undertaken by an individual in the age-group *j*. Thus, the total loss in value from stay-at-home for the group *(i, j)* is *f*_*ij*_*h*_*ij*_, unless the occupation *i* can be worked at home, which we allow in one of our specifications.

The goal is to find a policy *(V, H)* that minimizes the loss of lives and the economic burden from a stay-at-home order. In particular, the planner solves the following linear program:

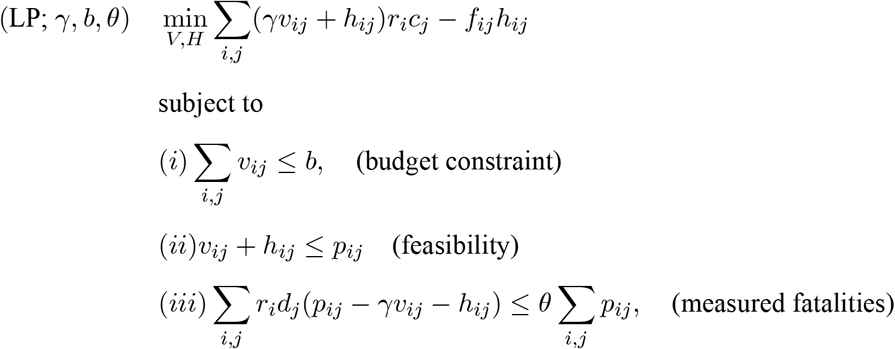

where *d*_*j*_ is the infection fatality rate rate that an individual in age group *j* faces. Constraint (*iii*) in this program allows us to derive the optimal policy *(V, H)* such that the (unconditional) fatality rate expected to occur in the population is capped, given a vaccine effectiveness of *γ*. Alternatively, constraint (*iii*) can inform us about the minimum vaccine effectiveness required to cap the (unconditional) fatality rate at *θ*, if a stay-at-home order is not possible. Depending on the values assumed for the parameter *θ*, constraint (*iii*) need not be binding.

## 3 Data and Estimation Strategy

We track 8 age-groups for the 2017 U.S. population, 16-19, 20-29, 30-39,…, and 80+, distributed over 454 occupations, aggregated at the 3-digit Census OCC code. We obtain the number of people for each age-group employed in a given occupation from the 2017 American Community Survey (ACS). Our sample is representative of 60% of the U.S. population.

To proxy for the benefit, *f*_*ij*_, that an individual in age group *j* generates from participating in economic activity by occupation *i*, we use the average yearly wage for each age group and occupation, also provided by the 2017 ACS. From an economics perspective, the wage captures a worker’s contribution to the production of total output as measured by GDP, or equivalently the GDP loss if a worker is unable to work due to a stay-at-home order (Hulten, 1978, Baqaee et al., 2020). We recognize that wages need not be a perfect proxy, with the value of some occupations for the economy potentially being underestimated. To overcome this limitation we designate certain occupations as essential. This is equivalent to assuming that workers in an essential occupation generate a very large value from participating in economic activity, regardless of their age.

The average cost, *c*_*j*_, for a person in age-group *j* that has been infected with the virus depends on the infection fatality rate, *d*_*j*_, that her age group faces and on the value of statistical life (VSL) for her age group. In particular, the cost of infection is given by

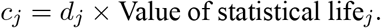

As Table 1 indicates, using VSL for each age-group is similar to using standard expected years of life lost (SEYLL), as Emanuel et al. (2020) suggests. For the infection fatality rate – the number of deceased among the infected people – we use the estimates provided by Salje et al. (2020) who jointly analyze French hospital data with the results of a detailed outbreak investigation aboard the Diamond Princess cruise ship.^2^ For the VSL, we use the estimates provided by Greenstone and Nigam (2020) who update the estimates of Murphy and Topel (2006) to 2015. The details are reported in Table 1.

**Table 1:** The Value of Statistical Life (VSL) and Infection Fatality Rate by age-groups. Note: for the age group 16-19, we used VSL of the ages 10-19.

| Age group | EU population | Infection fatality rate (%) | VSL in mil. USD | The cost of infection in USD |
| --- | --- | --- | --- | --- |
| < 19 | 0.215 | 0.001 | 15.3 | 153 |
| 20-29 | 0.12 | 0.005 | 16.1 | 805 |
| 30-39 | 0.135 | 0.02 | 15.8 | 3,160 |
| 40-49 | 0.14 | 0.05 | 13.8 | 6,900 |
| 50-59 | 0.135 | 0.2 | 10.3 | 20,600 |
| 60-69 | 0.115 | 0.7 | 6.7 | 46,900 |
| 70-79 | 0.09 | 1.9 | 3.7 | 70,300 |
| 80+ | 0.05 | 8.3 | 1.5 | 124,500 |

Note: for the age group 16-19, we used VSL of the ages 10-19.

The remaining variable that we need to estimate in the model is the infection rate, *r*_*i*_, associated with each occupation *i*, for which the data is not directly available. To circumvent the lack of data, we proceed in two steps. First, we infer the infection rate for each occupation group based on the coronavirus-related deaths by occupation that have occurred between March 9th and May 25th, 2020, as reported by the U.K. Office for National Statistics (ONS). ONS reports the age-standardized death rate per 100,000 of each minor occupation *i* by gender for those aged 20 to 64 years. This death rate is unconditional on infection and based on the 2013 E.U. standard population distribution. We use the employment-weighted average of the death rates by gender and construct the death rate, *D*_*i*_, per 100,000 people for each U.K. minor occupation. Given the infection fatality rate, *d*_*j*_, for age group *j* provided by Salje et al. (2020), we obtain the infection rate for each U.K. minor occupation per 100, 000 people as

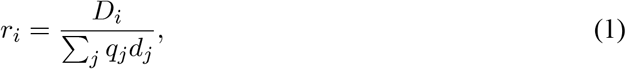

where *q*_*j*_ ϵ [0, 1] denotes the fraction of age-group *j* according to the E.U. standard population distribution.^3^ Our maintained assumption is that exposure to the virus depends on occupation, but the infection fatality rate depends on patients’ ages.

Next, we impute infection rates for the U.S. occupations. We consider that the main explanatory factor for differences in infection rates across occupations is physical proximity. In an occupation with a higher physical proximity score, workers have to interact more closely with other people, such as co-workers or clients. Thus, presumably, the virus transmission rate is higher in occupations that require a higher degree of physical proximity, and, consequently, this will be reflected in death rates. Even as various social distancing measures are observed, we expect that occupations with a higher degree of physical proximity will still entail a higher infection risk than ones with a lower degree of physical proximity.

We estimate a fractional probit model (Papke and Wooldridge, 1996) using the infection rates, *r*_*i*_, corresponding to each U.K. minor occupation we have derived based on (1) and physical proximity measures that are also provided by ONS. A worker employed in occupation *i* with degree of physical proximity *x*_*i*_ ϵ [0, 100] is going to be infected over two months with probability

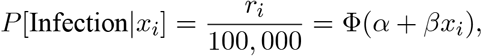

where *Φ* is the cumulative distribution function of the standard normal distribution. The estimates we obtain are 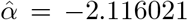 (CI [-2.592531, -1.639511]) and 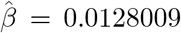 (CI [0.004813, 0.0207889]), which are statistically significant at 0.05 level. The resulting infection rates are thus increasing with the degree of physical proximity, ranging from 2,638 (for logging workers) to 20,160 (for physical therapists) per 100,000 people. Estimated infection rates over a two-month period for selected occupations are shown in Fig. 2.

**Figure 1:**
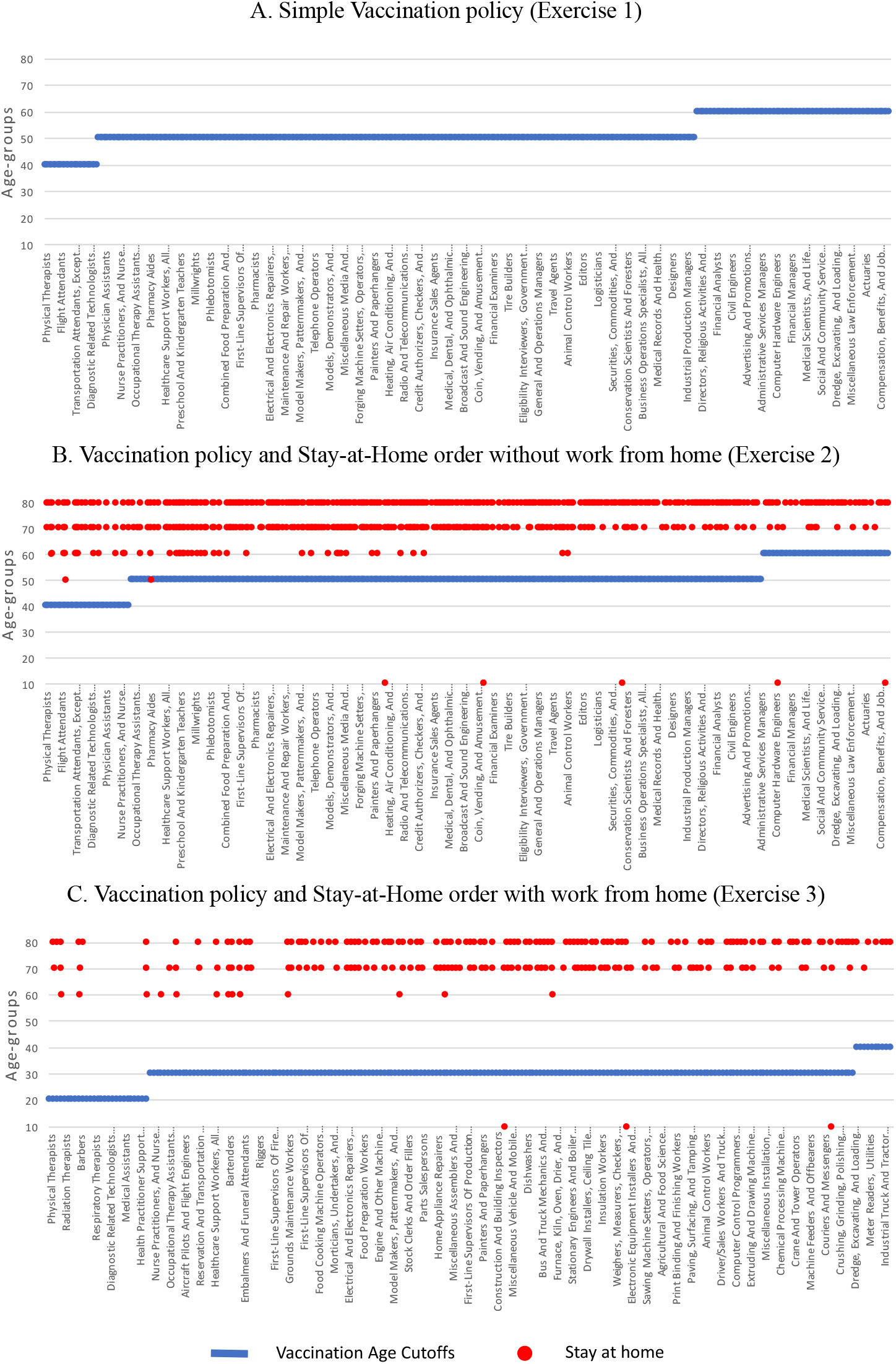
Age cutoffs for vaccinations and age groups staying at home. Occupations on the x-axis are ordered based on their infection risk. (A) The optimal vaccination policy showing the youngest age for each occupation that is eligible to receive the vaccine. (B) The optimal vaccination policy showing the youngest age for each occupation that is eligible to receive the vaccine, together with the occupation-age groups that are mandated to stay at home. (C) The optimal vaccination policy showing the youngest age for each occupation which cannot be done from home that is eligible to receive the vaccine, together with the occupation-age groups that are mandated to stay at home. Occupations that can be done from home do not receive a vaccine.

**Figure 2:**
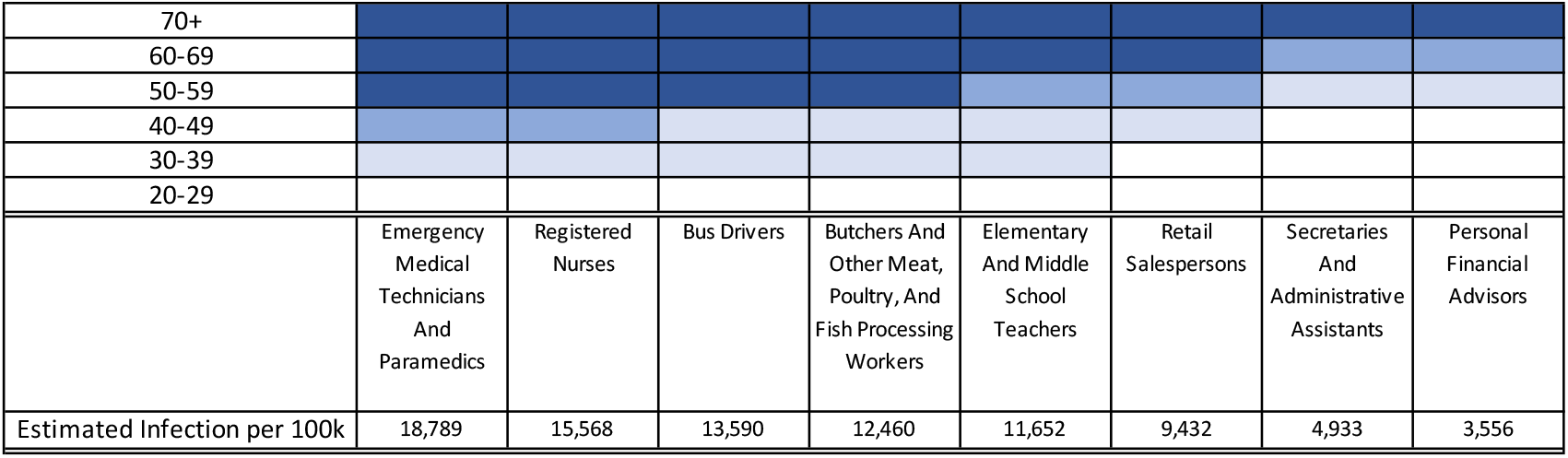
Priorities among some selected age-occupation groups. The groups with the top priority are marked in the darkest blue, and they receive vaccines even when the supply is 30 million doses. Lighter blues mark the groups that have the second and the third priority, and they will get vaccines when the supply is, respectively, 60 million and 100 million doses. The rest groups with the lowest priorities are marked in white.

We then impute the infection rate for each US occupation based on these estimates.^4^ In particular, we construct the infection rate per 100, 000 people over a two-month period for each U.S. occupation *i* with proximity score *x*_*i*_ ϵ [0, 100] as

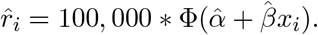

We use the proximity score developed by Mongey, Pilossoph, and Weinberg (2020) who calculate an the employment-weighted average of survey-based job characteristics for each 3-digit OCC occupation code based on O*NET data.

For robustness we estimate the model using mortality rates for ages 20+. For this, we calculate age-standardized mortality rates by occupation including the number of deaths of those aged 65+ that ONS provides for each occupation. In this case, the parameter estimates from the fractional probit regression are 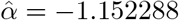 and 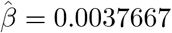. Including all ages yields a lower estimate of 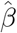 which implies that the infection rates estimated for U.S. occupations will be less responsive to proximity scores, i.e., more homogeneous infection rates across occupations. In this case, our main finding – vaccines to be distributed mostly by ages rather than occupations – becomes even stronger (Fig. A.5). Nevertheless, we are cautious about these estimates, as the death rate for the 65+ age category is bound to be less precise due to the small sample size of the elderly employed in each occupation.

## 4 Results

We undertake three exercises. First, we find the optimal vaccine distribution under the assumption that there is no stay-at-home order, and everyone returns to work regardless of whether they have received a vaccine or not. Second, we derive the optimal vaccine distribution when a targeted stay-at-home order is implemented and that the individuals who are unable to return to work produce no output. That is, everyone who cannot return to work receives no wage for the stay-at-home order duration. Third, we derive the optimal vaccine distribution when a targeted stay-at-home order is implemented, but for some occupations, employees can work from home. In this case, individuals that can work from home receive the same wages as if they were to return to work, while individuals that cannot work from home produce no output and receive no wages for the duration of the stay-at-home order. For the last two exercises, 121 occupations deemed to be essential are exempt from the stay-at-home order. We assume that the length of the stay-at-home order is 2 months (Abbasi, 2020) to reflect the expected time lag until a vaccine becomes widely available and scale the yearly wage loss accordingly. We also cautiously assume that the initial supply of the vaccine allocated to employed people above the age of 16 is 60mil doses, covering approximately one-third of the employed workforce. Similarly, we assume that the vaccine effectiveness is 50% (Food and Drug Administration, 2020). We initially derive the optimal vaccination and stay-at-home policy when the constraint on the fraction of coronavirus fatalities is lax.

The overall vaccine policy is presented in Fig. 1. We order occupations based on their infection risk, and show how the vaccine distribution policy and stay-at-home mandate depend on occupations and ages. The main insight from all three exercises is that age is more important than an occupation’s infection risk when allocating vaccines optimally. The largest volume of vaccines are allocated to the populations of age 50-59, followed by age 60-69 in exercise 1 and 2, or age 30-39 in exercise 3 (Table A.1). The loss in economic benefits (proxied by wages) from being out of work plays a role in allocating vaccines only when a stay-at-home order is also used as a policy tool (as in exercise 2 and 3).

When we derive the optimal vaccine allocation absent of a stay-at-home order, all employed people above age 60 receive the vaccine (Fig. 1A). Some occupations, such as paramedics and flight attendants, are eligible to receive the vaccine if they are at least 40 years old. For many other occupations, including most other healthcare workers, the eligibility threshold for receiving the vaccine is age 50. There is naturally a trade-off between the infection risk associated with occupation and the risk of death related to age. For instance, any food processing workers above age 50 receive a vaccine, while financial advisors only receive the vaccine if they are above 60 years old.

When a stay-at-home order complements the vaccination policy, most employees who are at least 80 years old, and some in their 70s are mandated to stay at home (Fig. 1B). For the 80+ age-group, the risk of death is so substantial that a 50% effective vaccine is insufficient to overcome the loss in wages for the duration of the stay-at-home order. For a few occupations such as textile-related, the stay-at-home order targets teenagers as well. While the infection fatality rate for their age group is meager, the economic value of practicing their occupation given the corresponding infection rate does not justify the risk. In turn, the stay-at-home order allows nurses as young as 40 years old to receive vaccines.

Once we take into account that for some occupations, employees can work from home without any loss in wages, then the supply of the vaccine can be distributed only towards those occupations in which employees need to be present at their workplace. Allocating vaccines across fewer occupations implies that younger people, for instance, as young as 20 for nurses and food preparation workers, are now eligible to receive the vaccine (Fig. 1C).

We illustrate how priorities are assigned across different age groups for some selected occupations in Fig. 2. The top priority groups are shaded in the darkest blue and the groups with the next priorities in lighter blues. The top priority groups consist of high-risk populations in high-exposure occupations, consistent with an emerging consensus. They receive vaccines even under a very limited supply (30 million doses). The following priority groups receive vaccines when the supply increases to 60 million doses or 100 million doses. Young healthcare workers such as paramedics and nurses at age 30+ (or 40+) have about the same priorities as financial advisors at age 50+ (or, respectively, 60+). A scarcer supply of the vaccine (30 million doses) emphasizes occupational risk, with nurses, for instance, being prioritized at age 50, while retail salespersons are eligible only at age 60.

Designating occupations as essential affect the optimal allocation of the vaccine only when a targeted stay-at-home order is also used (Fig. A.1). In designating, for instance, food processing workers as essential, we ensure that the individuals over 50 years old in this occupation receive vaccines. Otherwise, if food processing workers can be subject to the stay-at-home order, only the population under 70 years old, representing 99.3% of the workforce, can return to their workplace, with individuals over 50 years old (in exercise 2) or over 20 years old (in exercise 3) receiving the vaccine.

Finally, our model allows us to derive the proportion of people that will still be infected with the virus even under the optimal vaccination policy, given any effectiveness and supply of the vaccine. When the vaccine is 50% effective, 7.14% of the employed workforce will still get the virus over the two months until the vaccine becomes widely available. When some occupations can be done at home, the proportion decreases to 2.69%. An increase in the vaccine effectiveness to 70% does not change the vaccine allocation (Fig. A.2), but reduces the fraction of infected people close to 6.5% (exercise 1 and 2) or to 2.07% (exercise 3). However, the fraction of infected people decreases also as the supply of the vaccine increases. With 100mil doses, not only that the age of the youngest eligible recipients decreases (Fig. A.4), but also the fraction of infected people declines to 6.15% (in exercise 1) and 5.92% (in exercise 2) even with vaccine effectiveness of 50%.

One may find that even the optimal vaccination policy would surrender to too many infections and potentially too many fatalities. For the (unconditional) mortality rate to be comparable with the one from the average flu, the vaccine effectiveness should be at least 65% and a stringent stay-at-home order targeting all but essential occupations that cannot be done from home is necessary, provided the supply is 60mil doses.^5^ Without the stringent stay-at-home order, a vaccine must be at least 91% effective and supplied in at least 150mil doses to reduce the fatalities of SARS-CoV-2 to the level of an average influenza season.

## 5 Conclusions

A model in which the population faces a death risk that depends on age and an infection risk that depends on occupation allows us to determine the optimal vaccine distribution policy for all U.S. employed population above the age of 16. Identifying priority groups for COVID-19 vaccination is critical for implementation planning, and our analysis can be input into how to allocate the vaccine across different populations.

In deriving the optimal allocation of vaccines based on ages and occupations, we considered exposures to the coronavirus through occupations. Naturally, people can also get exposed while spending time with family, shopping, or engaging in leisurely activities. These exposures outside of occupations suggest that the optimal vaccine allocation should be tilted even more towards the elderly. In other words, exposure to infection risk outside working hours dampens the occupational priorities in the vaccine allocation.

## Data Availability

All data is public and available through U.K. ONS or U.S. Census Bureau.

## A Supplementary Text

### A.1 Physical Proximity Scores

O*NET asks a number of questions about individuals’ working conditions and day-to-day tasks of their job. To evaluate proximity, the question asks, “How physically close to other people are you when you perform your current job?”. Respondents provide a response on a scale between one and five, one indicating that the respondent does not work near other people (beyond 100ft.), while five indicating that they are very close to others (near touching). More information on these questions is provided in the Instructions for Work Context Questionnaire (Q 21), published by O*NET.

The responses to this question are standardized by Mongey, Pilossoph, and Weinberg (2020)to a scale ranging from 0 to 100 as follows. First, O*NET reports the answers to the survey using the fine occupation SOC-code. Mongey, Pilossoph, and Weinberg (2020) calculate an employment-weighted average, 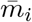 of the response to this question that corresponds to each occupation, *i*, classified according to the 3-digit Census OCC code. Second, Mongey, Pilossoph, and Weinberg (2020) follow the procedure used by O*NET and re-scale the survey answer to the interval [0, 100] using the following equation:

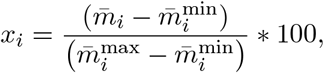

where *x*_*i*_ is the final physical proximity standardized score for occupation *i*.

### A.2 Work-from-Home Occupations

The work-from-home classification of occupations has been developed by Mongey, Pilossoph, and Weinberg (2020) and Dingel and Neiman (2020) using O*NET survey answers from the Work Activities Questionnaire and Work Context Questionnaire. The Work Context Questionnaire includes questions aiming to capture the “physical and social factors that influence the nature of work” such as interpersonal relationships, physical work conditions, and structural job characteristics. The Work Activities Questionnaire includes questions aiming to capture the “general types of job behaviors occurring on multiple jobs” such as the input of information, mental processes, and work output.

We use the classification of Mongey, Pilossoph, and Weinberg (2020) because they provide an employment weighted aggregation at the 3-digit Census OCC codes. In particular, they aggregate 18 occupational attributes based on answers to the questions Q4, Q14, Q17, Q18, Q29, Q33, Q37, Q43, Q44, in the Work Context Questionnaire, and Q4A, Q16A, Q17A, Q18A, Q20A, Q22A, Q23A, Q32A in The Work Activities Questionnaire. The responses to these questions are standardized to a work-from-home score ranging from 0 to 1 following the same procedure as the one used to calculate the physical proximity score. In the next step, an occupation is classified either as that it can be done from home if its work-from-home score is above the median, or that it cannot be done from home if its work-from-home score is below the median.

We use this classification to derive the optimal allocation of vaccines when a stay-at-home order is used, and some occupations can be done from home (exercise 3). However, we acknowledge that this classification has limitations, as some occupation categories may be too coarse. For instance, physicians and surgeons have been classified as a work-from-home occupation. While for many physicians telemedicine seems feasible for limited periods of time, as has been evident during the lockdown in the U.S., we understand that telemedicine is not applicable for surgeons or critical care doctors. Another example is teachers, who are also classified as a work-from-home occupation. At the same time, our approach is flexible, and an optimal vaccine allocation can be derived under various specifications, including fractional ones, for which occupations are done from home. Thus, as some physicians return to hospitals, and some teachers return to teach in person, these occupations can be re-classified partly as occupations that are not done from home, and our model will assign vaccines accordingly.

### A.3 Essential Occupations

We have designated occupations to be essential based on the guidelines issued by the Cybersecurity and Infrastructure Security Agency (CISA). Our classification is inherently subjective and is also subject to the limitation that some occupations are very coarse. For instance, we have classified network and computer systems administrators as an essential occupation according to the guidelines issued by CISA. However, we acknowledge that it is likely that not all system administrators are essential workers. It is re-assuring that designating an occupation as essential plays no role when a simple vaccination policy is considered, as in exercise 1.

**Table A.1:** Vaccine Distribution by Ages.

|  | 20-29 | 30-39 | 40-49 | 50-59 | 60-69 | 70-79 | 80+ |
| --- | --- | --- | --- | --- | --- | --- | --- |
| Exercise 1 | 0 | 0 | 2,518,118 | 26,165,823 | 24,040,335 | 6,094,810 | 1,180,914 |
| Exercise 2 | 0 | 0 | 3,664,572 | 30,441,166 | 21,867,803 | 3,659,431 | 367,028 |
| Exercise 3 | 2,994,988 | 16,111,831 | 15,109,864 | 15,223,264 | 8,768,288 | 1,567,305 | 224,460 |

**Table A.2:** The Infection Fatality Rate by age-groups reported in South Korea.

| Age group | Infection fatality rate (%) |
| --- | --- |
| < 19 | 0 |
| 20-29 | 0 |
| 30-39 | 0.08 |
| 40-49 | 0.14 |
| 50-59 | 0.41 |
| 60-69 | 1.29 |
| 70-79 | 6.32 |
| 80+ | 20.05 |

**Figure A.1:**
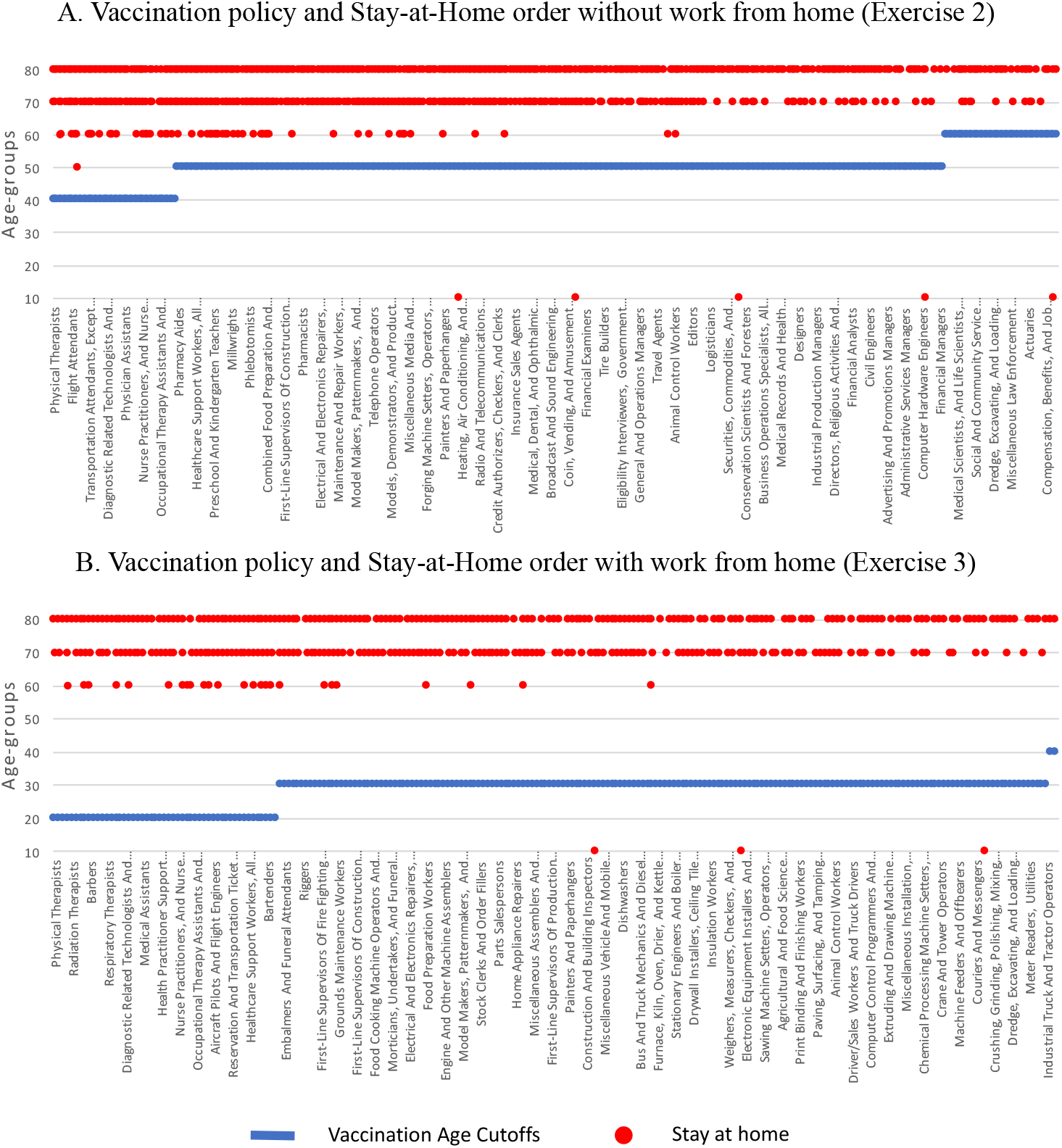
Age cutoffs for vaccinations and age groups staying at home when no occupations are designated to be essential. Occupations on the x-axis are ordered based on their infection risk. (A) The optimal vaccination policy showing the youngest age for each occupation that is eligible to receive the vaccine, together with the occupation-age groups that are mandated to stay at home. (B) The optimal vaccination policy showing the youngest age for each occupation, which cannot be done from home that is eligible to receive the vaccine, together with the occupation-age groups that are mandated to stay at home. Occupations that can be done from home do not receive a vaccine. Note: We omit exercise 1, which remains the same as in Fig. 1A. Exercise 1 requires workers of all occupations to return to work regardless of being vaccinated or not. Designating an occupation to be essential does not affect the optimal vaccine allocation in this case.

**Figure A.2:**
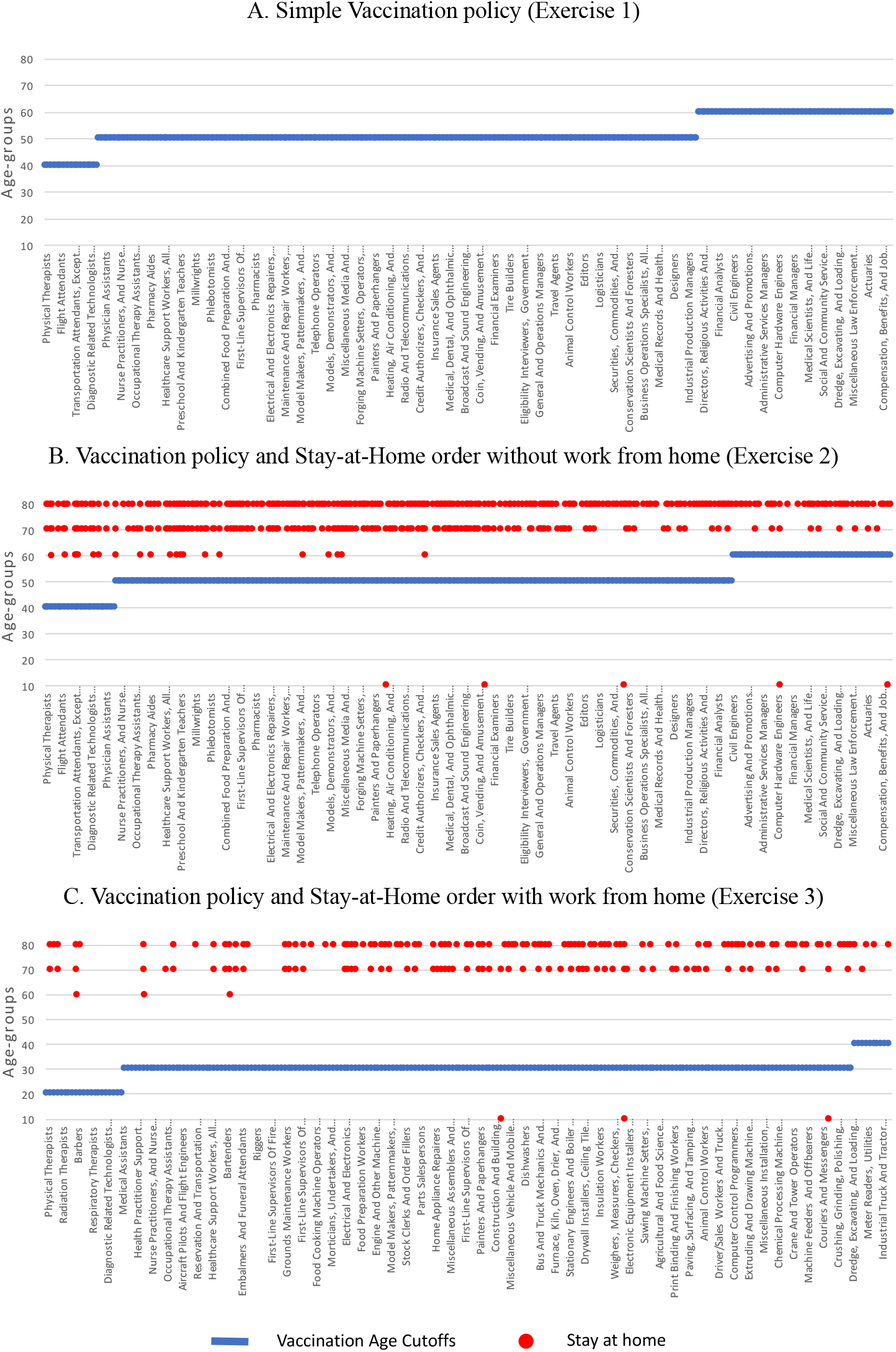
Age cutoffs for vaccinations and age groups staying at home when vaccine effectiveness is 0.7. Occupations on the x-axis are ordered based on their infection risk. (A) The optimal vaccination policy showing the youngest age for each occupation that is eligible to receive the vaccine. (B) The optimal vaccination policy showing the youngest age for each occupation that is eligible to receive the vaccine, together with the occupation-age groups that are mandated to stay at home. (C) The optimal vaccination policy showing the youngest age for each occupation which cannot be done from home that is eligible to receive the vaccine, together with the occupation-age groups that are mandated to stay at home. Occupations that can be done from home do not receive a vaccine.

**Figure A.3:**
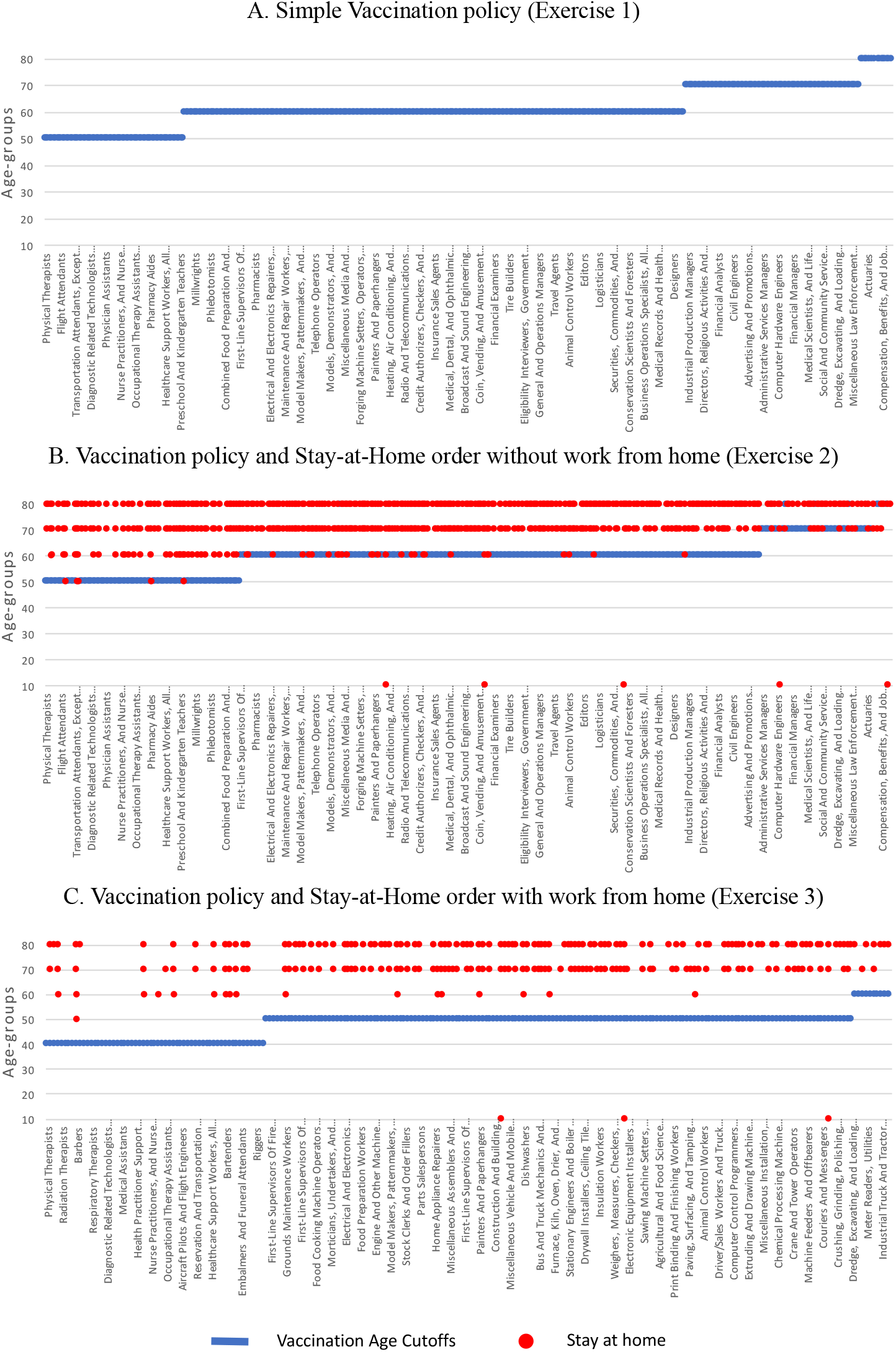
Age cutoffs for vaccinations and age groups staying at home, when the supply of vaccines is 30mil doses. Occupations on the x-axis are ordered based on their infection risk. (A) The optimal vaccination policy showing the youngest age for each occupation that is eligible to receive the vaccine. (B) The optimal vaccination policy showing the youngest age for each occupation that is eligible to receive the vaccine, together with the occupation-age groups that are mandated to stay at home. (C) The optimal vaccination policy showing the youngest age for each occupation which cannot be done from home that is eligible to receive the vaccine, together with the occupation-age groups that are mandated to stay at home. Occupations that can be done from home do not receive a vaccine.

**Figure A.4:**
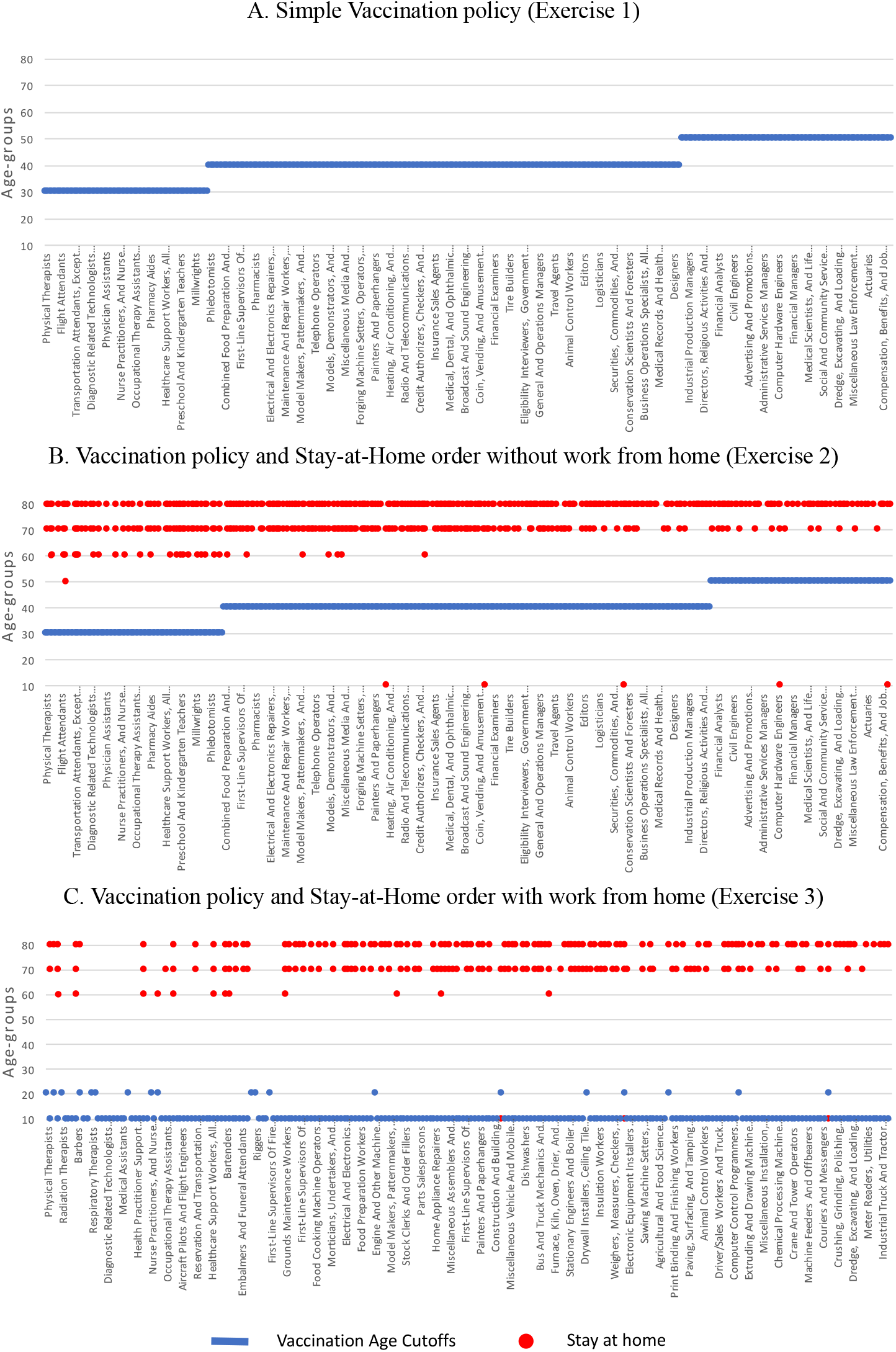
Age cutoffs for vaccinations and age groups staying at home, when the supply of vaccines is 100mil doses. Occupations on the x-axis are ordered based on their infection risk. (A) The optimal vaccination policy showing the youngest age for each occupation that is eligible to receive the vaccine. (B) The optimal vaccination policy showing the youngest age for each occupation that is eligible to receive the vaccine, together with the occupation-age groups that are mandated to stay at home. (C) The optimal vaccination policy showing the youngest age for each occupation which cannot be done from home that is eligible to receive the vaccine, together with the occupation-age groups that are mandated to stay at home. Occupations that can be done from home do not receive a vaccine. Note: While, in exercise 3, the vaccination cutoffs may appear to be non-monotonic in occupations’ risks, that is only because some occupations have no teenage workers.

**Figure A.5:**
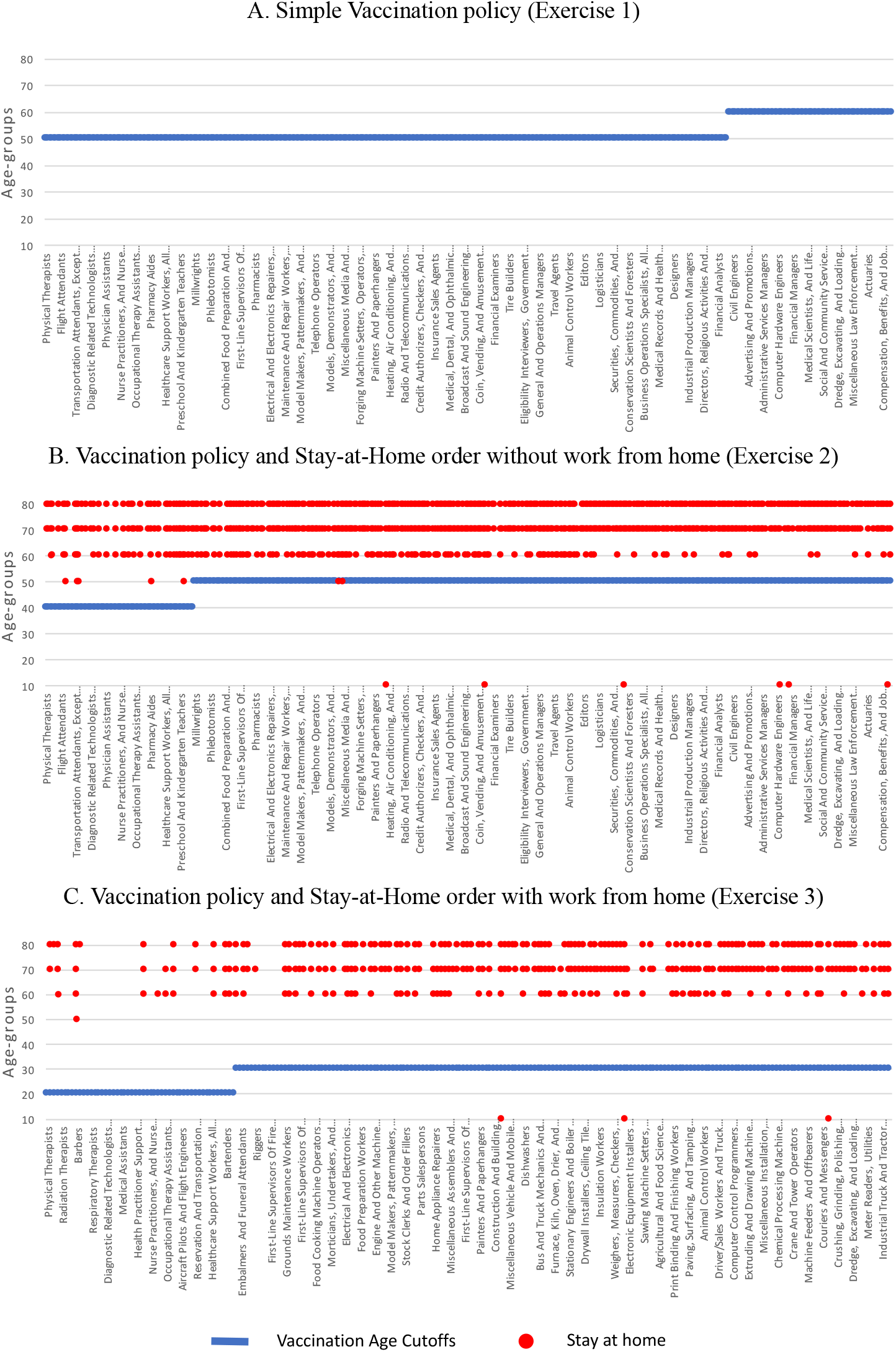
Age cutoffs for vaccinations and age groups staying at home (probit estimation including all ages mortality risk, *α*=-1.152288 and (*β*=0.0037667). Occupations on the x-axis are ordered based on their infection risk. (A) The optimal vaccination policy showing the youngest age for each occupation that is eligible to receive the vaccine. (B) The optimal vaccination policy showing the youngest age for each occupation that is eligible to receive the vaccine, together with the occupation-age groups that are mandated to stay at home. (C) The optimal vaccination policy showing the youngest age for each occupation which cannot be done from home that is eligible to receive the vaccine, together with the occupation-age groups that are mandated to stay at home. Occupations that can be done from home do not receive a vaccine.

**Figure A.6:**
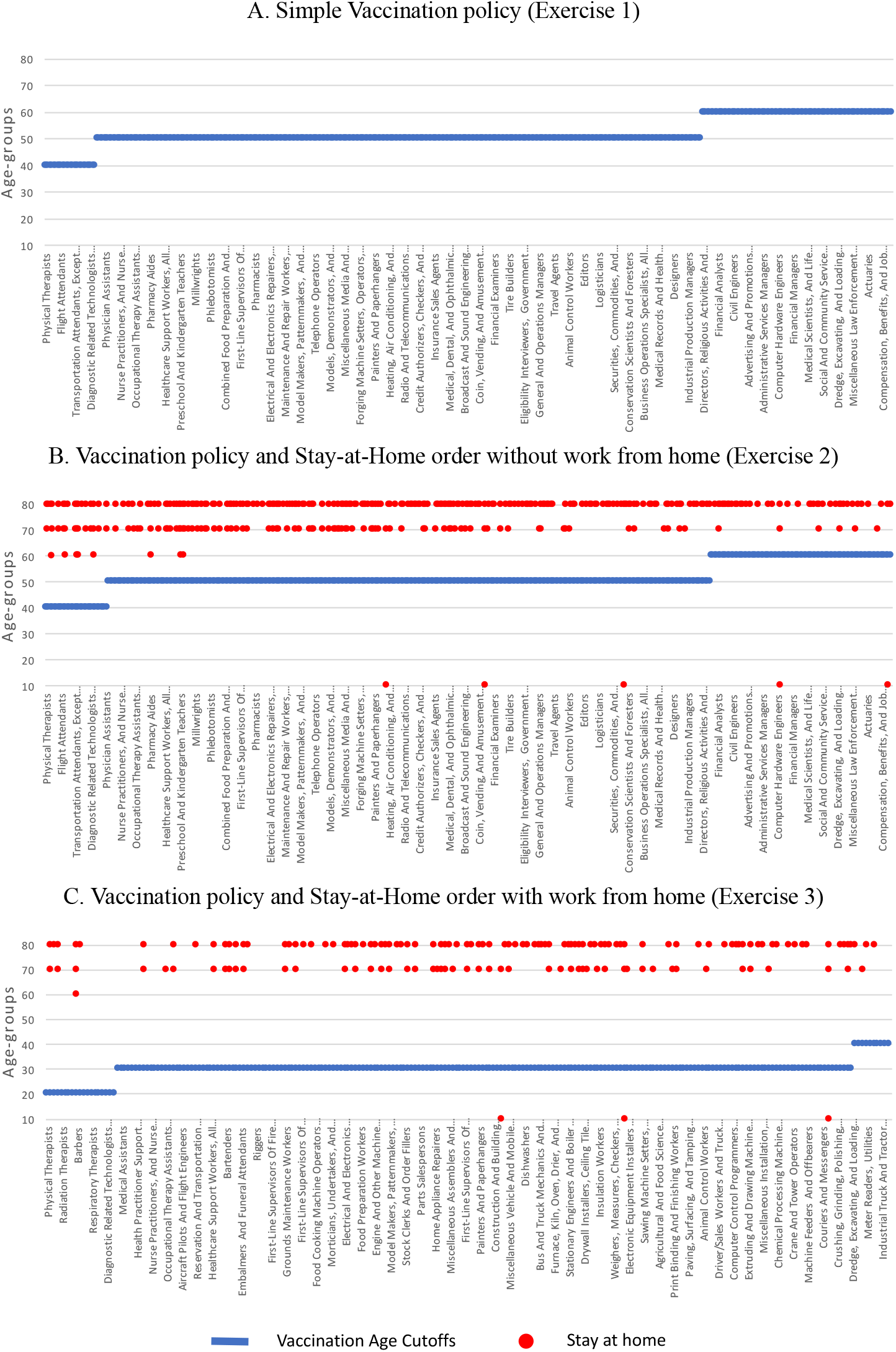
Age cutoffs for vaccinations and age groups staying at home based on South-Korean infection fatality rates from Table A.2. Occupations on the x-axis are ordered based on their infection risk. (A) The optimal vaccination policy showing the youngest age for each occupation that is eligible to receive the vaccine. (B) The optimal vaccination policy showing the youngest age for each occupation that is eligible to receive the vaccine, together with the occupation-age groups that are mandated to stay at home. (C) The optimal vaccination policy showing the youngest age for each occupation which cannot be done from home that is eligible to receive the vaccine, together with the occupation-age groups that are mandated to stay at home. Occupations that can be done from home do not receive a vaccine.

## Footnotes

1 The implementation is not trivial as some healthcare workers are also elderly. For example, an elderly-first implementation favors, ironically, healthcare workers. Elderly healthcare workers receive vaccines based on their age rather than on their occupations, and this way extra vaccines are available to younger healthcare workers. Hence, healthcare workers can receive vaccines in excess of the doses reserved for them.

2 In a separate robustness exercise (Fig. A.6 and Table A.2) we also use the reported infection fatality rate data from South Korea, which has a very accurate track-and-trace system. Although mortality rates in South Korea are different than the ones estimated in France, the relative mortality risk across different age groups shows a similar patern. As such, the optimal vaccination policy remains largely the same.

3 Since the death *D*_*i*_ is based on those aged 20 to 64 years, we calculate infection rates using the fraction, *q*_*j*_, of age-group *j* ϵ {(20 - 29), (30 - 39), (40 - 49), (50 - 59), (60 - 64)} out of the total population aged 20 to 64 years.

4 One may consider bypassing the infection rate by matching the U.K. death rates by occupation to U.S. occupations. We do not take such approach as we need to find unconditional death rate for each occupation *and* age-group.

5 CDC reports that seasonal flu has resulted in between 12,000 - 61,000 deaths annually since 2010. We take the average 38,000 and divide it by 6 to account for a two-month period in our setup. The resulting number of flu-related deaths represents about 0.012% of about 320 million U.S. population.

